# Smartphone-Based Assessment of Cognitive Variability in Alzheimer Disease

**DOI:** 10.64898/2026.11.22.26368632

**Authors:** Andrew Jin Soo Byun, Ko Woon Kim, Meaghan McKenna, John Torous, Chun Lim

## Abstract

**INTRODUCTION:** Repeated smartphone testing can measure cognitive inconsistency that a single clinic visit cannot capture. We evaluated cognitive variability and everyday phone use in biomarker-confirmed Alzheimer disease (AD).

**METHODS:** Older adults with mild cognitive impairment due to AD or mild AD dementia and cognitively unimpaired controls used *Smartphone Monitoring Assessment in Real Time–Alzheimer (SMART-A)* on their own phones. Response-time variability was adjusted for task demand and practice.

**RESULTS:** Among 26 participants with cognitive data (14 controls, 12 AD), the four-marker variability composite was higher in AD (Hedges g = 1.29; 95% confidence interval, 0.53–2.06; P = .003) and correlated with baseline Montreal Cognitive Assessment score (r = −0.60; P = .001). Memory-task accuracy was lower in AD at all four trials, and 64.5% versus 98.2% of sessions reached delayed recall (P < .001). Median phone-use bouts were shorter in AD (2.4 vs 5.4 minutes; P = .031), without a detectable pickup-frequency difference. Diary-reported bedtime was earlier in AD; sensor-derived sleep, activity, and mobility showed no statistically detectable differences. None of the 19 cross-domain contrasts survived false-discovery-rate correction.

**DISCUSSION:** SMART-A supported repeated assessment over months in biomarker-confirmed AD. Cognitive variability, memory-task progression, and phone-use duration are promising complementary measures, requiring independent validation of reliability, incremental value, and sensitivity to clinical change.

**Highlights:**

- Repeated smartphone testing captured greater cognitive instability in biomarker-confirmed AD.
- Greater cognitive instability was associated with poorer clinical cognitive performance.
- AD participants showed lower memory accuracy and less progression to delayed recall.
- Shorter everyday phone-use bouts provided an exploratory behavioral signal.
- Findings support validation of smartphone measures to complement clinic-based assessment.

**Research in context:** *Systematic review:* We examined published studies of intraindividual variability, unsupervised digital cognition, and passive monitoring. Prior work supports cognitive variability as a candidate indicator of impairment, but its clinical relevance during extended home testing in biomarker-confirmed symptomatic AD remains less established.

*Interpretation:* SMART-A captured greater cognitive instability associated with poorer clinical cognitive performance, alongside reduced memory-task progression and exploratory differences in everyday phone use. These findings suggest that repeated home assessment may characterize aspects of cognitive impairment beyond a single clinic visit; incremental clinical value remains unproven.

*Future directions:* Independent studies should establish whether these measures improve detection or monitoring beyond mean task performance and conventional cognitive assessments. Prespecified endpoints and harmonized observation windows will be essential to determine whether within-person changes track clinically meaningful cognitive or functional decline.

## INTRODUCTION

Conventional cognitive assessments characterize impairment at a clinical visit but provide limited information about how consistently an individual performs across everyday occasions. This distinction is relevant to Alzheimer disease (AD), in which emerging treatments have increased the need for accessible measures that can characterize cognitive impairment and support longitudinal assessment [1,2]. Individuals with similar average performance may differ in the frequency and magnitude of their cognitive fluctuations. Measuring these fluctuations could therefore provide a complementary perspective on cognitive function that is not captured by a single assessment.

Intraindividual variability (IIV) refers to fluctuations in an individual’s cognitive performance across repeated observations. Greater response-time IIV has been associated with cognitive impairment, AD biomarkers, and subsequent cognitive outcomes, supporting its investigation as a potential marker of cognitive vulnerability [3,4,5]. Response-time distributions can also reveal unusually slow responses that are obscured by average scores [6]. However, observed variability reflects several influences, including overall response speed, task demands, and practice [7]. Evaluating IIV therefore requires repeated, detailed measurements that allow these influences to be considered when interpreting performance inconsistency.

Smartphone-based cognitive assessment provides an opportunity to obtain these measurements outside the clinic. Previous studies have demonstrated the feasibility and validity of unsupervised digital testing, detected greater cognitive variability in mild cognitive impairment, and identified amyloid-related differences in learning during repeated assessment [8,9,10,11]. Using mixed-effects location scale models, Aschenbrenner and colleagues found greater session-to-session processing-speed variability in cognitively normal APOE ε4 carriers during one week of smartphone testing [12]. Nevertheless, the characterization of cognitive variability during extended home assessment in symptomatic, biomarker-confirmed AD remains less established. In particular, it remains unclear whether repeated testing over months can identify performance inconsistency associated with AD after accounting for practice and task demands. Addressing this gap would help establish whether variability is a useful candidate measure for further clinical validation.

SMART-A (Smartphone Monitoring Assessment in Real Time–Alzheimer) is well suited to investigating this question because it combines repeated cognitive testing on participants’ own smartphones with records of response timing, accuracy, and task progression [13]. Its DigitSpan, TicTacToe Memory, and Trails tasks provide repeated observations across working memory, episodic memory, and executive function. These records enable examination of within-person response-time variability while accounting for repeated exposure and recorded task demands. Extended assessment also makes it possible to study learning and task completion alongside variability, providing a broader characterization of performance than scores from completed tests alone. In parallel, SMART-A collects routine phone-use measures, sleep reports, and passive sensor data, allowing cognitive findings to be examined alongside everyday behavioral patterns [13,14,15,16].

Following the published protocol [13], we studied community-dwelling older adults with biomarker-confirmed mild cognitive impairment due to AD or mild AD dementia and cognitively unimpaired controls. Our primary objective was to determine whether response-time variability and a composite incorporating variability and unusually slow responses differed between groups after accounting for task demand and practice. We also examined the composite’s association with conventional cognitive performance. Exploratory analyses evaluated memory-task progression, everyday phone use, and diary- and sensor-derived sleep measures. Together, these analyses assessed the feasibility and potential clinical relevance of extended smartphone measurement as a foundation for subsequent validation.

## METHODS

### Participants

Community-dwelling older adults were recruited through a memory-disorders clinic and by community referral under the study protocol [13]. The AD group comprised mild cognitive impairment due to AD and mild AD dementia using current criteria [17]. Controls had no cognitive diagnosis.

All 13 participants in the AD group had a biomarker-confirmed diagnosis [17]. Eight patients had a positive amyloid-PET scan (21.1–148.5 Centiloid), 5 patients had plasma p-tau217 above the upper cut point (range 0.47–1.23), and two patients had positive CSF. Two patients had both amyloid-PET and ptau217 testing. The Beth Israel Deaconess Medical Center institutional review board approved the study (2024P000931), and all participants provided written informed consent.

Thirty-two people were screened and 28 were eligible. Not everyone contributed every kind of data, so each analysis states its own sample: 26 (14 control, 12 AD) had enough cognitive sessions, 26 (13 control, 13 AD) had screen-use data, 24 had both sensor and diary sleep, and 19 had usable location data. We never pooled across these. All 27 appear in Table 1.

**Table 1.**
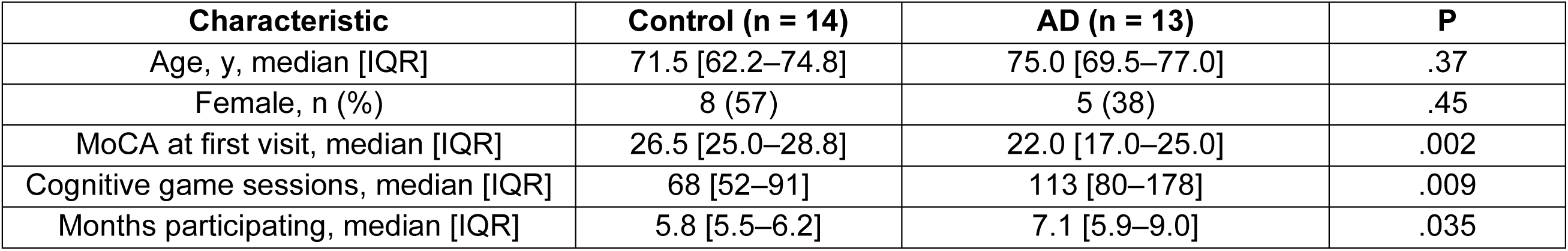
Cohort characteristics and engagement.

### The app and the cognitive tasks

Participants used SMART-A, built on the mindLAMP platform [18], on their own phones, at their own pace, for several months. Three short games were delivered automatically on a weekly basis as well as on demand. DigitSpan showed a string of digits to repeat back, first in the order given and then in reverse, with the string growing longer until the participant made two errors at one length. TicTacToe Memory showed three picture-and-location pairs, repeated them three times so they could be learned, then asked for them again after a delay. Trails asked the participant to tap numbers and letters in alternating order.

### Task-record interpretation

The app records tap-level response times and accuracy, together with a trial or stage identifier. These identifiers have different meanings across tasks and do not always represent comparable levels of difficulty. We therefore applied task-specific restrictions to ensure interpretable group comparisons.

In DigitSpan, trials are numbered consecutively from 1 to 17 across the forward and backward recall blocks, while digit-sequence length resets at the transition to backward recall. Because participants may switch blocks at different points, the same trial number can represent different sequence lengths and recall directions. For example, it may correspond to an eight-digit forward sequence for one participant and a three-digit backward sequence for another. Group comparisons were therefore restricted to trials 1–2, which consistently assess forward recall.

In TicTacToe Memory, trial number identifies the phase of the memory assessment: trials 1–3 represent repeated learning of the same picture–location pairs, and trial 4 assesses delayed recall. Each trial was analyzed separately to distinguish performance during learning from performance at delayed recall.

In Trails, each session comprises three sequential stages containing 12, 20, and 20 targets, respectively. Although participants progress through these stages, individual tap records are retained only for the highest stage reached. Consequently, the recorded stage reflects progression through the task rather than an independently assigned difficulty level. Analyses were restricted to stage 3 and therefore describe performance only in sessions that progressed beyond stages 1–2. These results should be interpreted as performance among sessions reaching the final stage, rather than as overall Trails performance across all sessions.

### Measuring variability

Response times lengthen with task difficulty and shorten with repeated exposure; both sources of systematic variation inflate apparent inconsistency. Variability was therefore estimated from model residuals rather than from raw response times. For each task, log-transformed response time was modelled on centered session index (practice) and centered trial level (task demand), with per-participant random effects fitted by restricted maximum likelihood: a random intercept and a random slope for trial level where level varied within a session, and a random intercept alone for Trails, which was analyzed at a single stage. Intraindividual variability was defined as the within-person standard deviation of these residuals, the dispersion that remains once practice and task demand have been accounted for. This adjustment removes two known influences on variability but does not render the measure independent of mean response speed.

Tap-level response times at Trails stage 3 were additionally fitted with an ex-Gaussian distribution, whose τ parameter indexes the slow tail; estimates were restricted to participants contributing at least 50 taps. The composite instability index was the mean of four standardized markers: residual intraindividual variability from DigitSpan, TicTacToe Memory and Trails, together with Trails τ. Each marker was oriented so that higher values denote greater instability, and the index was computed for participants with at least three of the four. It therefore combines variability across repeated administrations with a distributional feature of individual response times.

### Sleep

Each morning participants reported what time they fell asleep and woke up. A weekly questionnaire administered the PROMIS Sleep-Related Impairment short form [19]. We kept diary nights reporting 3 to 14 hours in bed, which retained 3,264 of 3,709 records with the same retention in both groups. Sleep was estimated from phone sensors using a hidden Markov model that classified half-hour intervals as sleep or wake [20].

### Phone use and passive sensors

A phone-use episode, or bout, ran from unlocking the phone to the next lock, capped at four hours. We measured how long bouts lasted, total daily screen time, the share of bouts lasting more than ten minutes, and how many times a day the phone was picked up. Call length came from the phone’s call records; for 6 of 27 participants that field contained timestamps rather than durations and those records were excluded. Movement and location measures covered activity level, day-to-day variability, fragmentation, circadian rhythm, time spent at home and distance travelled, with all timestamps converted to each participant’s own local time.

### Statistical analysis

We summarize each group difference as Hedges’ g, which expresses the gap between the two group averages in units of how spread out the individual scores are. Because the groups are small, g is adjusted downward slightly to correct a known upward bias [21]. For each g we also report the range of values that remain consistent with the data, obtained by re-drawing the sample with replacement 10,000 times and reading off the middle 95% of the resulting estimates.

Group comparisons used two-sided Welch t-tests; participant characteristics were compared with two-sided Mann–Whitney or Fisher exact tests. Pearson correlation described the association between the composite and MoCA [22]. Benjamini–Hochberg q values are reported for the 19 cross-domain comparisons in Figure 6. Sample sizes differ between data streams, and comparisons of effect size across domains are descriptive.

## RESULTS

### Participant characteristics and engagement

Table 1 describes the 27 participants contributing to at least one analysis. MoCA was lower in AD (median 22.0 vs 26.5; P = .002), while age and sex comparisons were not statistically significant. Participants with AD completed more cognitive sessions (median 113 vs 68; P = .009) over longer participation (7.1 vs 5.8 months; P = .035).

### Task performance

On the TicTacToe memory task, accuracy was lower in AD at every one of the four trials (g = −1.30, −1.63, −1.62, −1.81; P = .006, .002, .002, .001), and responses were slower at trials 2 to 4 (Figure 2). On DigitSpan, accuracy was lower in AD at the first forward trial (g = −0.98, P = .021); the difference at the second forward trial was not statistically significant (g = −0.38, P = .32), while how far participants got did not differ (P = .77). On Trails, responses were slower in AD at stage 3 (g = 1.02, P = .021). AD sessions reached the delayed-memory probe in 64.5% of cases versus 98.2% in controls (P < .001; Figure 2c).

**Figure 1.**
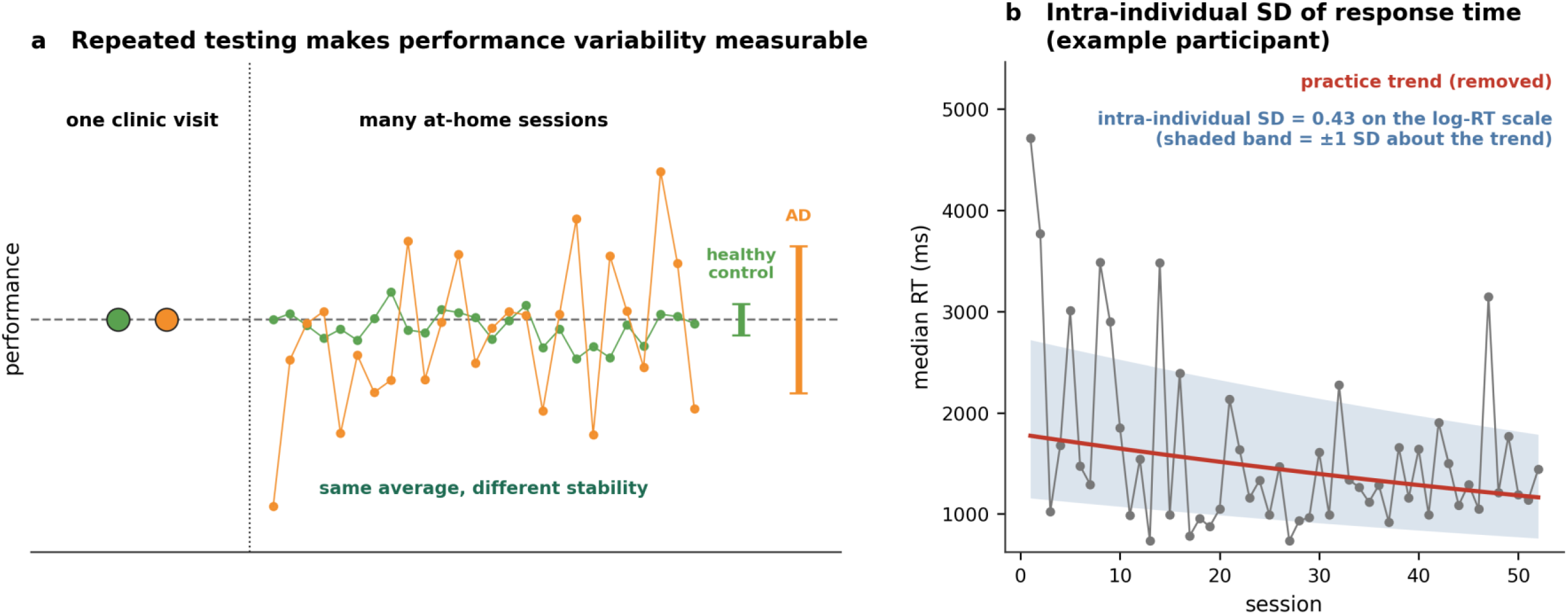
Repeated assessment of cognitive variability. (a) Schematic of two individuals with equivalent mean performance and different session-to-session consistency. (b) Response-time series for one participant, shown with the fitted practice trend. Shaded band, ±1 SD of the residuals about the trend; the annotated value is on the log response-time scale.

**Figure 2.**
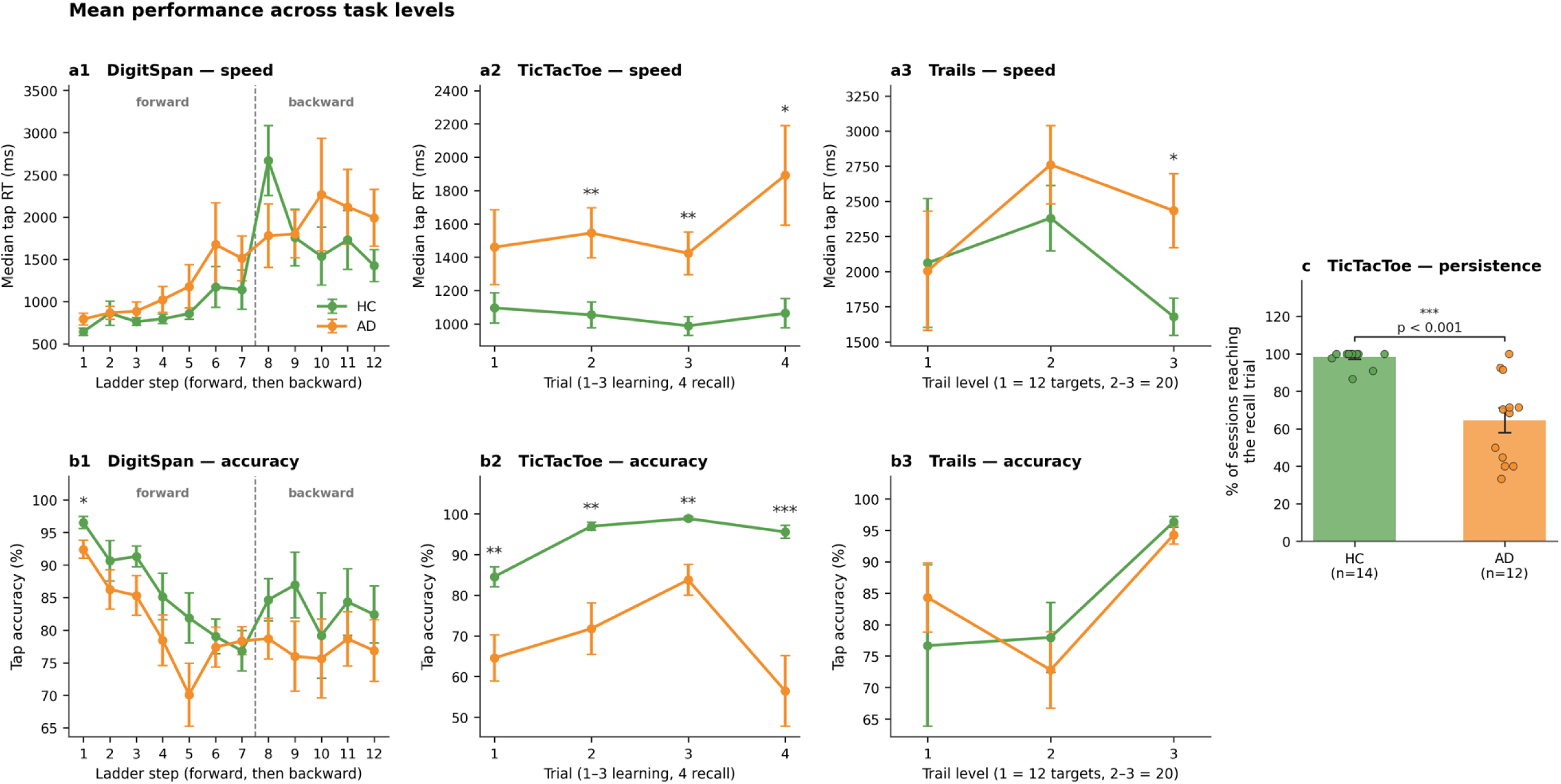
Performance across task trials. Points are group means ± SEM of per-participant values: median tap response time (a1–a3) and mean tap accuracy (b1–b3) at each level. Level definitions differ by task (Table 2). (c) Percentage of TicTacToe Memory sessions reaching the delayed-recall trial. Welch t-test: *P < .05, **P < .01, ***P < .001. n = 14 control, 12 AD in all panels.

**Table 2.** Cognitive task description and analysis approach.

| **Task** | **Session structure** | **Meaning of recorded level** | **Analysis approach** |
|---|---|---|---|
| DigitSpan | Forward digit recall followed by backward digit recall. Sequence length resets when the task switches from forward to backward recall. | Sequential trial number (1–17), which continues across the switch in recall direction. | Group comparisons were restricted to trials 1–2, which consistently assess forward recall. |
| TicTacToe Memory | Three picture–location pairs are presented across three learning trials, followed by delayed recall. | Position in the task sequence: learning trials 1–3 and delayed recall at trial 4. | Group comparisons were conducted separately for each of the four trials. |
| Trails | Three sequential stages comprising 12, 20, and 20 alternating targets, respectively. | Highest stage reached during the session. Individual tap records are retained only for that stage. | Analyses were restricted to stage 3; results therefore reflect performance among participants who progressed beyond stages 1–2. |

### Intraindividual variability

After accounting for trial difficulty and practice, the combined inconsistency score was higher in AD (14 control, 12 AD; g = 1.29; 95% CI, 0.53-2.06; P = .003; Figure 3a), and it was also significantly correlated with the MoCA at the first visit (r = -0.60; P = .001; Figure 3e).

**Figure 3.**
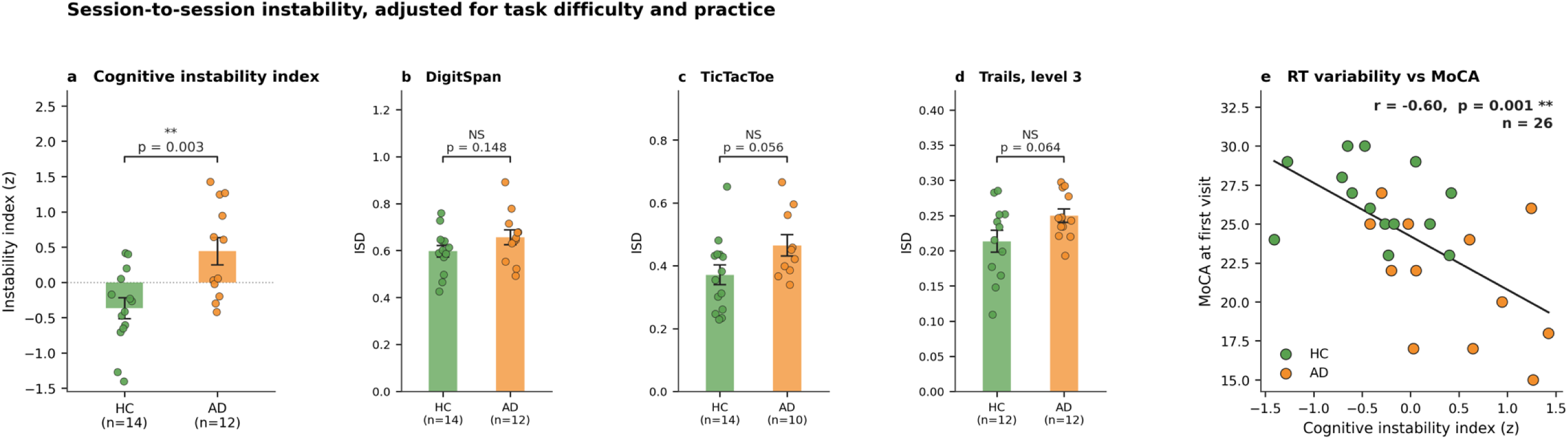
Cognitive variability adjusted for task difficulty and practice. (a) Composite of four standardized markers. (b–d) Residual intraindividual variability by task. (e) Composite against MoCA at the first visit. Bars, group mean ± SEM; points, individual participants. Sample sizes are given on each axis. Welch t-test in (a–d); Pearson correlation in (e).

Trails τ showed the largest individual variability contrast (g = 1.22; P = .007). Improvement in DigitSpan accuracy across sessions was smaller in AD (g = −0.92; P = .021). The individual residual-IIV estimates were higher in AD but did not reach the p=.05 threshold (TicTacToe Memory g = 0.80; Trails g = 0.78; DigitSpan g = 0.58). The composite had a larger observed standardized contrast than these residual-IIV components.

### Smartphone usage pattern

Episodes of ordinary phone use were about half as long in AD (median 2.4 vs 5.4 minutes; g = 0.91, P = .031), mean daily screen time was lower (150 vs 299 minutes; g = 0.94, P = .021), and a smaller mean share of bouts lasted beyond ten minutes (18.2% vs 33.7%; g = 1.00, P = .017). Calls were shorter in the same direction, though not significantly (g = 0.88, P = .099). The mean number of times the phone was picked up each day did not differ (23.5 vs 26.6; P = .56). Screen-use analyses included 13 controls and 13 participants with AD; call analyses, 9 and 12 (Figure 5). Study activities (cognitive games) overlapped 2.4% of phone-use bouts. Excluding those bouts yielded a similar duration contrast (g = 0.95; P = .030), whereas bouts containing study activities showed no detectable difference (P = .64).

**Figure 4.**
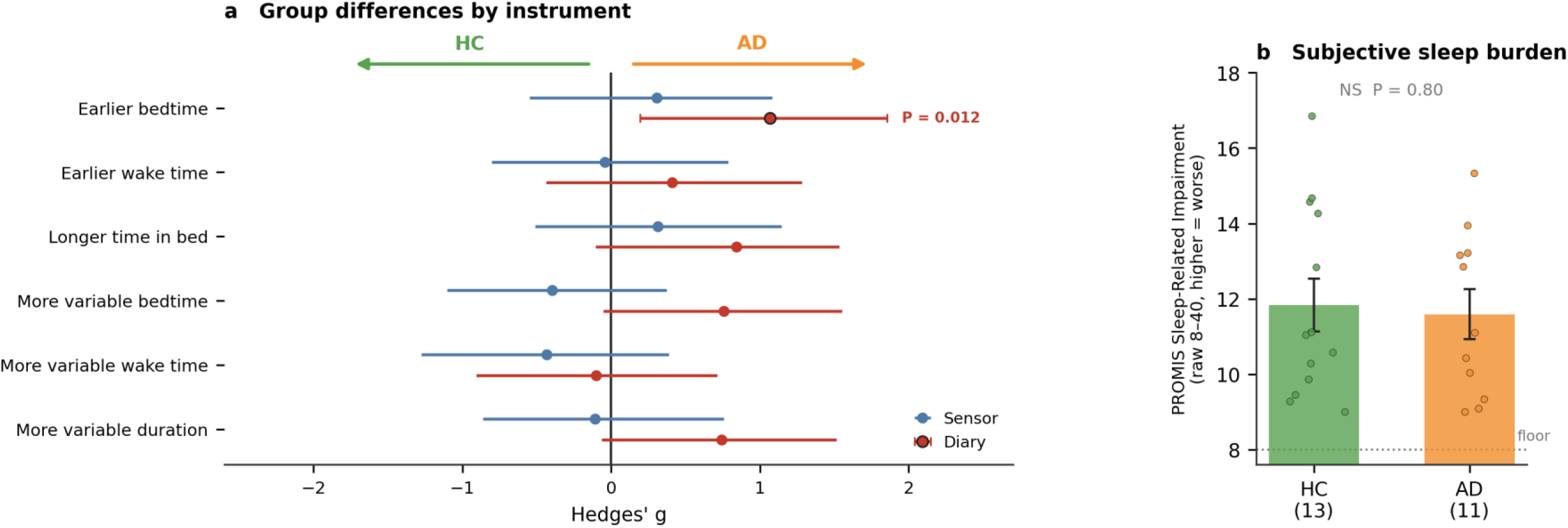
Diary and sensor-derived sleep measures (n = 24). (a) Standardized group contrasts for matched measures: Hedges’ g with 95% bias-corrected and accelerated bootstrap confidence intervals. (b) PROMIS Sleep-Related Impairment, raw score (range 8–40; higher indicates greater impairment). Welch t-test.

**Figure 5.**
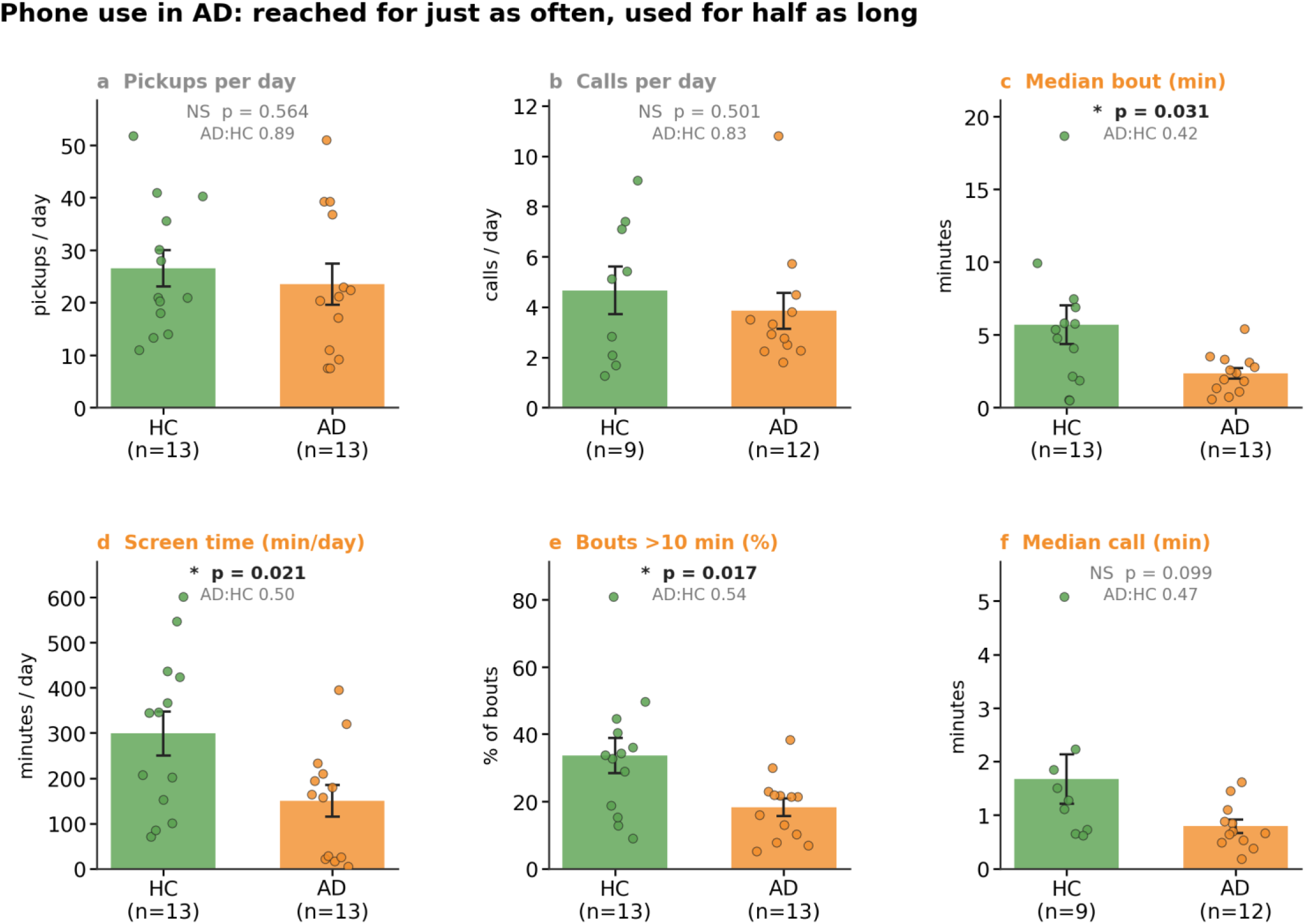
Everyday phone use. Daily pickups, daily calls, median bout duration, daily screen time, proportion of bouts exceeding ten minutes, and median call duration. Bars, group mean ± SEM; points, individual participants. AD:HC, ratio of group means. Screen-use panels, 13 control and 13 AD; call panels, 9 and 12. Welch t-test.

**Figure 6.**
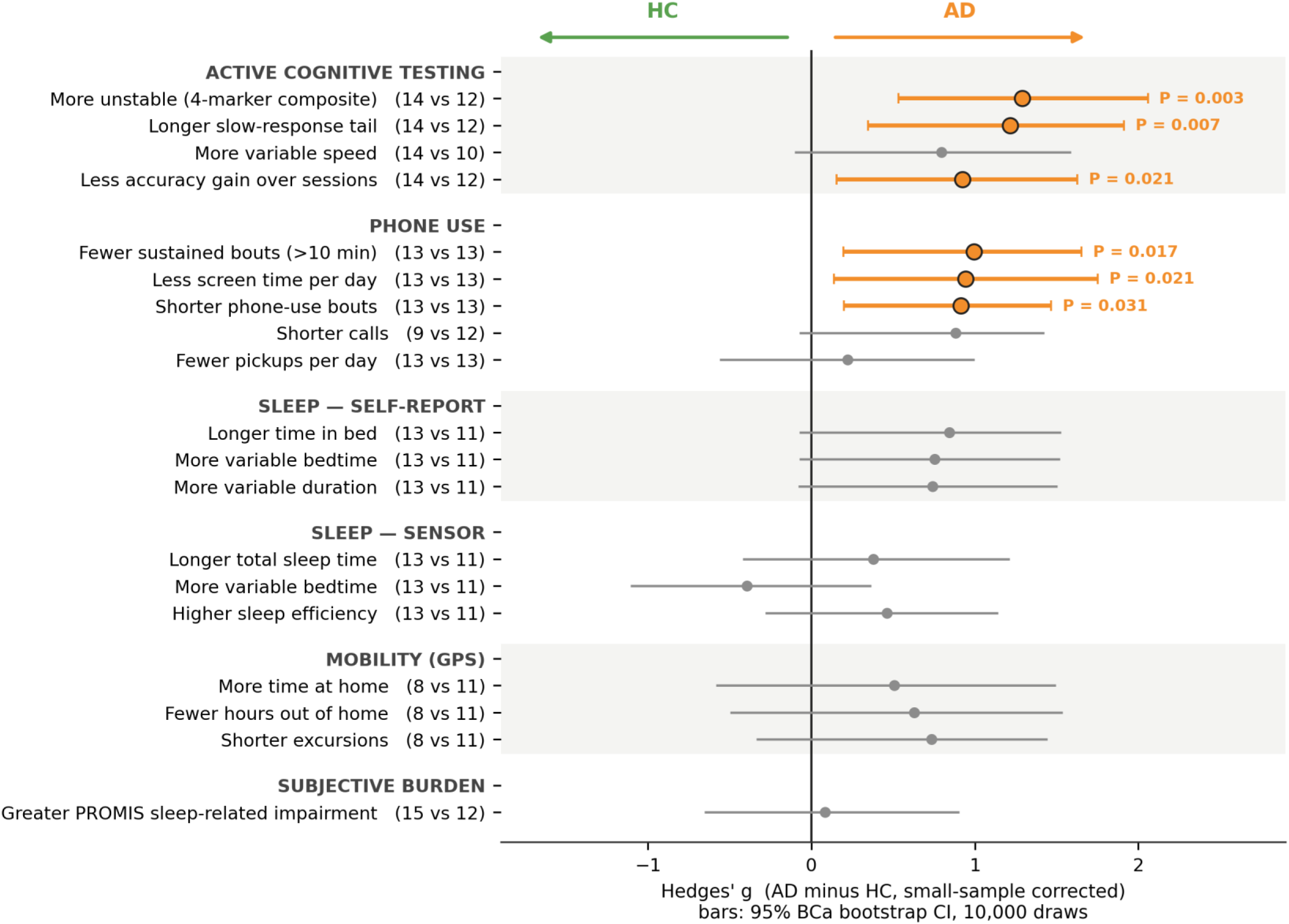
Cross-domain comparison of group differences. Points, Hedges’ g (AD minus control, small-sample corrected); bars, 95% bias-corrected and accelerated bootstrap confidence intervals from 10,000 resamples. Rows are oriented so that rightward indicates the direction characteristic of AD. Color denotes intervals excluding zero. Sample sizes are given per row as (control vs AD). No comparison survived Benjamini–Hochberg correction across the 19 contrasts (smallest q = .06).

### Self-reported and sensor-derived sleep

Among the 24 participants with both measures, diary reports showed earlier bedtimes in the AD group (g = −1.11, P = .012); longer time in bed (g = 0.84, P = .062) and greater night-to-night variability in bedtime (g = 0.75, P = .071) (Figure 4). Time in bed (g = 0.74, P = .079) were in the same direction but did not reach the nominal threshold. Sensor-derived estimates showed no statistically detectable group differences (all P ≥ .23). PROMIS Sleep-Related Impairment did not differ between groups (11.8 vs 11.6; P = .80), with scores in both groups near the floor of the scale, indicating little reported sleep-related impairment.

### Cross-domain comparison

Figure 6 summarizes 19 cross-domain contrasts. Six met the nominal threshold: the composite, Trails τ, DigitSpan learning, and three phone-use measures. Paired diary bedtime differed in the separate sleep analysis (P = .012). No cross-domain contrast survived false-discovery-rate correction (smallest q = .06).

## DISCUSSION

Repeated assessment with SMART-A revealed a pattern of cognitive and behavioral differences between older adults with symptomatic AD and cognitively unimpaired controls. The cognitive instability composite was higher in AD and associated with lower MoCA scores, while memory accuracy, progression to delayed recall, and everyday phone-use duration also differed between groups. Participants with AD contributed a median of 113 cognitive sessions over 7.1 months, demonstrating that substantial repeated measurements can be obtained during extended home assessment. These findings support an approach to digital assessment that characterizes the consistency and unfolding of cognitive performance alongside conventional measures of speed and accuracy.

The higher instability composite is broadly concordant with laboratory evidence linking cognitive impairment to increased response-time variability [3,7]. Similarly, the COMPASS-ND study found greater reaction-time inconsistency in AD than in cognitively unimpaired, subjective cognitive impairment, and mild cognitive impairment groups [23]. However, our findings refine this interpretation: the clearest individual variability-related difference concerned the slow tail of the Trails response-time distribution, whereas residual IIV measures showed positive but statistically nonsignificant group differences. This pattern is particularly consistent with Tse and colleagues, who found that very mild AD disproportionately affected the slow tail of response-time distributions across attention tasks [6]. Our findings suggest that unusually slow responses remain informative during unsupervised smartphone testing. Schumacher and colleagues also reported an increased slow-response tail in AD using ex-Gaussian analysis of attention-task performance [24]. Such responses could reflect intermittent difficulty maintaining task goals or coordinating cognitive operations, although interruptions and other features of home testing provide alternative explanations.

The absence of statistically significant differences in individual residual-IIV measures also requires consideration. Prior findings are not uniform across tasks or measurement timescales: the Einstein Aging Study identified greater within-day variability in MCI for processing speed and visual memory binding, but not spatial working memory [10]. Aschenbrenner and colleagues likewise found greater session-level variability in APOE ε4 carriers for processing speed, but not consistently across working-memory and episodic-memory tasks [12]. Our mixed task-level findings are therefore compatible with domain-dependent sensitivity rather than a uniform increase in variability across cognition. Differences in task structure, self-selected testing occasions, and the longer observation period may also contribute. Importantly, Phillips and colleagues showed that adjustment for mean response speed eliminated an IIV difference in MCI but not AD [7]. Although our models accounted for practice and recorded task demand, they did not establish independence from average speed. Aschenbrenner and colleagues’ location scale models jointly estimated mean performance and variability; our residual-based approach differs and does not establish the same separation between mean performance and variability [12].

SMART-A extends existing mobile assessment approaches through sustained measurement in symptomatic disease. ARC established the feasibility and validity of brief smartphone assessments administered repeatedly over seven days, and the Einstein Aging Study demonstrated the sensitivity of ambulatory variability measures during a 16-day assessment period [8,10]. Our study complements these intensive assessment designs by examining self-paced participation over several months.

The association between the instability composite and MoCA provides preliminary evidence that smartphone-derived inconsistency relates to clinically recognizable cognitive impairment. This is consistent with the broader construct-validity evidence supporting remote cognitive assessment [8]. Nevertheless, the pooled correlation may partly reflect separation between diagnostic groups, and it does not establish that instability captures impairment beyond mean task performance or conventional testing.

Memory-task findings provide both convergent evidence and an additional perspective on remote assessment. Lower TicTacToe Memory accuracy across learning and delayed-recall trials agrees with evidence that unsupervised digital memory tests can detect AD-related impairment [9]. Reduced improvement in DigitSpan accuracy across sessions is also conceptually consistent with studies showing diminished learning during repeated digital testing in amyloid-positive cognitively unimpaired adults [11], although the tasks, disease stages, and learning intervals differ. Together, these findings support examining the trajectory of repeated performance as well as its average level. SMART-A additionally captured whether sessions reached delayed recall: only 64.5% of AD sessions did so, compared with 98.2% of control sessions. This difference highlights task progression as a potentially informative assessment feature and a source of selection in analyses restricted to completed trials.

The phone-use findings extend the assessment beyond explicit cognitive tasks. Participants with AD had shorter phone-use bouts and lower daily screen time, despite no statistically detectable difference in pickup frequency. The duration contrast persisted after excluding bouts containing study activities, suggesting that it reflected ordinary device use rather than simply the cognitive testing schedule. Large multimodal studies have demonstrated the potential of combining interactive assessments with smartphone and wearable behavior to identify cognitive impairment [14]. We seek to identify a readily interpretable candidate feature within this broader approach: the duration of engagement after phone use begins. Shorter bouts could reflect difficulty sustaining complex interactions, but application preferences, social opportunities, habitual routines, and automatic locking settings could produce similar patterns.

Sleep findings were less consistent across measurement approaches. Diaries indicated earlier bedtime in AD, whereas phone-derived sleep estimates showed no statistically detectable group differences. These results do not necessarily contradict studies associating altered rest–activity rhythms with dementia [15,16]. Those studies examined wearable-derived rhythms and prospective dementia outcomes, whereas our analyses compared phone-derived estimates between small groups with established diagnostic differences. A phone records device movement and interaction, which may diverge from the participant’s sleep or activity when the device is unattended. Diary reports introduce different uncertainties, including recall and reporting error.

Taken together, these findings advance the field by demonstrating how one smartphone platform can support extended assessment of performance consistency, learning, task progression, and everyday engagement in symptomatic AD. Detailed response records make it possible to account for systematic changes associated with practice and task demand, while parallel behavioral monitoring generates hypotheses about functioning outside formal testing. This creates a practical foundation for investigating whether changes in cognitive performance and everyday behavior converge within individuals over time.

Several limitations constrain interpretation. The small number of independent participants limits precision and generalizability, despite the substantial number of repeated sessions. None of the 19 cross-domain comparisons survived false-discovery-rate correction, so the findings require independent replication with prespecified outcomes. Unequal observation periods and potentially informative missingness may influence group contrasts. The composite combines residual variability with a response-distribution parameter and should not be interpreted as a pure measure of day-to-day fluctuation. Future studies should also assess reliability, harmonize observation windows, and test whether these measures add information beyond mean speed, accuracy, and baseline clinical cognition.

SMART-A demonstrates that extended home assessment can yield clinically relevant candidate measures of cognitive consistency and everyday engagement in older adults with symptomatic AD. Its principal contribution is a way to examine how cognition is expressed repeatedly in daily life, including variability, learning, and incomplete task progression. Independent validation could establish whether these measurements provide a useful bridge between episodic clinic assessments and the evolving cognitive and functional difficulties experienced by patients.

## Data Availability

All analyses were performed in Python and scripts written can be shared upon reasonable request.

## Funding

This study was funded by the a2 Pilot Awards, a subaward of funding given to the Trustees of the University of Pennsylvania by the National Institute on Aging of the National Institutes of Health under Award Number P30AG073105.

## Ethics

Approved by the Beth Israel Deaconess Medical Center institutional review board (2024P000931). All participants gave written informed consent.

## Use of artificial intelligence

Claude (Anthropic) and ChatGPT (OpenAI) were used to assist with language editing, manuscript restructuring, and reference checking. The authors are responsible for reviewing and verifying the final text, citations, and interpretation.

## Data and code availability

All analyses were performed in Python and scripts written can be shared upon reasonable request.

## Notes

### Competing Interest Statement

The authors have declared no competing interest.

### Author Declarations

The Beth Israel Deaconess Medical Center institutional review board approved the study (2024P000931), and all participants provided written informed consent.

### Summary of Updates

Previous version didn't include all others authors so just updated the author list

## REFERENCES

1. van Dyck CH, Swanson CJ, Aisen P, et al. Lecanemab in early Alzheimer’s disease. N Engl J Med. 2023;388(1):9–21.

2. Sims JR, Zimmer JA, Evans CD, et al. Donanemab in early symptomatic Alzheimer disease: the TRAILBLAZER-ALZ 2 randomized clinical trial. JAMA. 2023;330(6):512–527.

3. Gorus E, De Raedt R, Lambert M, Lemper JC, Mets T. Reaction times and performance variability in normal aging, mild cognitive impairment, and Alzheimer’s disease. J Geriatr Psychiatry Neurol. 2008;21(3):204–218.

4. Duchek JM, Balota DA, Tse CS, Holtzman DM, Fagan AM, Goate AM. The utility of intraindividual variability in selective attention tasks as an early marker for Alzheimer’s disease. Neuropsychology. 2009;23(6):746–758.

5. Bielak AAM, Hultsch DF, Strauss E, MacDonald SWS, Hunter MA. Intraindividual variability in reaction time predicts cognitive outcomes 5 years later. Neuropsychology. 2010;24(6):731–741.

6. Tse CS, Balota DA, Yap MJ, Duchek JM, McCabe DP. Effects of healthy aging and early stage dementia of the Alzheimer’s type on components of response time distributions in three attention tasks. Neuropsychology. 2010;24(3):300–315.

7. Phillips M, Rogers P, Haworth J, Bayer A, Tales A. Intra-individual reaction time variability in mild cognitive impairment and Alzheimer’s disease: gender, processing load and speed factors. PLoS One. 2013;8(6):e65712.

8. Nicosia J, Aschenbrenner AJ, Balota DA, et al. Unsupervised high-frequency smartphone-based cognitive assessments are reliable, valid, and feasible in older adults at risk for Alzheimer’s disease. J Int Neuropsychol Soc. 2023;29(5):459–471. doi:10.1017/S135561772200042X.

9. Berron D, Olsson E, Andersson F, et al. Remote and unsupervised digital memory assessments can reliably detect cognitive impairment in Alzheimer’s disease. Alzheimers Dement. 2024;20(7):4775–4791. doi:10.1002/alz.13919.

10. Cerino ES, Katz MJ, Wang C, et al. Variability in cognitive performance on mobile devices is sensitive to mild cognitive impairment: results from the Einstein Aging Study. Front Digit Health. 2021;3:758031. doi:10.3389/fdgth.2021.758031.

11. Papp KV, Jutten RJ, Soberanes D, et al. Early detection of amyloid-related changes in memory among cognitively unimpaired older adults with daily digital testing. Ann Neurol. 2024;95(3):507–517. doi:10.1002/ana.26833.

12. Aschenbrenner AJ, Hassenstab J, Morris JC, Cruchaga C, Jackson JJ. Relationships between hourly cognitive variability and risk of Alzheimer’s disease revealed with mixed-effects location scale models. Neuropsychology. 2024;38(1):69–80. doi:10.1037/neu0000905.

13. McKenna M, Torous J, Rozenblit E, Flathers M, Ryan S, Byun AJS, Lim C. Real-time smartphone monitoring assessments as a cognitive biomarker of Alzheimer disease: protocol for a development study. JMIR Res Protoc. 2026;15:e93259. doi:10.2196/93259.

14. Butler PM, Yang J, Brown R, et al. Smartwatch- and smartphone-based remote assessment of brain health and detection of mild cognitive impairment. Nat Med. 2025;31(3):829–839. doi:10.1038/s41591-024-03475-9.

15. Liu Y, Feng H, Du J, Yang L, Xue H, Zhang J, et al. Associations between accelerometer-measured circadian rest-activity rhythm, brain structural and genetic mechanisms, and dementia. Psychiatry Clin Neurosci. 2024;78(7):393–404. doi:10.1111/pcn.13671.

16. Wang W, Wanigatunga AA, Etzkorn LH, et al. Association between circadian rest-activity rhythms and incident dementia in older adults: the Atherosclerosis Risk in Communities Study. Neurology. 2026;106(2):e214513. doi:10.1212/WNL.0000000000214513.

17. Jack CR Jr, Andrews JS, Beach TG, et al. Revised criteria for diagnosis and staging of Alzheimer’s disease: Alzheimer’s Association Workgroup. Alzheimers Dement. 2024;20(8):5143–5169.

18. Torous J, Wisniewski H, Bird B, et al. Creating a digital health smartphone app and digital phenotyping platform for mental health and diverse healthcare needs. J Technol Behav Sci. 2019;4(2):73–85.

19. Buysse DJ, Yu L, Moul DE, et al. Development and validation of patient-reported outcome measures for sleep disturbance and sleep-related impairments. Sleep. 2010;33(6):781–792.

20. Byun AJS, Li Y, Cong S, Dhima A, Wang S, Flathers M, Torous J. Sleep estimation from low frequency smartphone sensors via Bayesian hidden Markov model. Research Square [preprint]. 2025. doi:10.21203/rs.3.rs-7217304/v1.

21. Hedges LV. Distribution theory for Glass’s estimator of effect size and related estimators. J Educ Stat. 1981;6(2):107–128.

22. Nasreddine ZS, Phillips NA, Bedirian V, et al. The Montreal Cognitive Assessment, MoCA: a brief screening tool for mild cognitive impairment. J Am Geriatr Soc. 2005;53(4):695–699.

23. Caballero HS, McFall GP, Gee M, et al. Cognitive speed in neurodegenerative disease: comparing mean rate and inconsistency within and across the Alzheimer’s and Lewy body spectra in the COMPASS-ND study. J Alzheimers Dis. 2024;100(2):579–601.

24. Schumacher J, Cromarty R, Gallagher P, et al. Structural correlates of attention dysfunction in Lewy body dementia and Alzheimer’s disease: an ex-Gaussian analysis. J Neurol. 2019;266(7):1716–1726. doi:10.1007/s00415-019-09323-y.

